# Developing a Global Framework for Digital Health in Traumatic Brain Injury (TBI): Clinician Perspectives of the Use of Digital Technologies in the TBI Care Pathway

**DOI:** 10.64898/2026.07.17.26358327

**Authors:** O Mantle, B G Smith, C Whiffin, L Hobbs, V Penmetcha, A Menon, S Venturini, T Bashford, P J Hutchinson

## Abstract

**Background:** Traumatic brain injury (TBI) affects 69 million individuals globally each year, yet care remains fragmented across complex, multi-specialty pathways and settings. Digital health technologies offer potential to bridge care gaps, particularly in resource-limited settings, yet existing frameworks do not adequately address the complexities of the TBI care pathway or the diverse global contexts in which care occurs.

**Methods:** A cross-sectional qualitative study using critical realist-informed thematic analysis was conducted with practising neurosurgeons recruited internationally via National Institute for Health and Care Global Health Research Group on Acquired Brain and Spine Injury (NIHR ABSI) collaborating centres, social media, and society newsletters. Semi-structured interviews were conducted by a single researcher (OM) via Microsoft Teams (March-July 2024), exploring technology availability, healthcare infrastructure, clinical pathways, and contextual challenges, with a systems thinking approach guiding identification of current and potential technology integration points. Fourteen neurosurgeons from twelve countries participated, representing six lower-middle, two upper-middle, and four high-income countries.

**Results:** Six inductive themes emerged: Availability, Acceptability, Applicability, Capability, Feasibility, and Possibility- forming a novel conceptual framework visualised as a hexagonal chart for guiding digital health technology design and implementation in TBI care. Marked disparities in technology availability and utilisation were identified across urban/rural settings and income levels.

**Conclusions:** This framework offers a practical, context-sensitive tool for researchers, policymakers, and clinicians developing or implementing digital health technologies in TBI care globally. Visualisation in a similar style to a radar-chart enables simultaneous consideration of factors- including digital literacy, infrastructure, and cultural attitudes- whose neglect frequently underlies implementation failures.

**Author Summary:** Traumatic brain injury affects millions of people every year, yet the care patients receive after their injury is often fragmented and inconsistent- particularly in lower-income countries where resources vary in quantity and accessibility. Digital technologies, from smartphone apps to telemedicine platforms, have the potential to bridge these gaps, but there is little practical guidance on how to design and implement them in ways that work across different healthcare settings around the world.

We interviewed neurosurgeons from twelve countries across three income settings to understand how digital technologies are currently used in traumatic brain injury care, and where they could make the biggest difference. We found that access to technology, attitudes towards it, the practicalities of healthcare systems, and the specific challenges of different settings all shape whether digital tools succeed or fail in practice.

From these conversations, a new framework, presented like a hexagonal radar chart, was developed to help researchers, clinicians, and policymakers think through the key factors that determine whether a digital health technology will work in each context. This tool supports more thoughtful, context-sensitive technology development and implementation in traumatic brain injury care worldwide, ultimately improving outcomes for patients regardless of where they live.

## Introduction

Traumatic brain injury (TBI) is a leading cause of death and disability worldwide, affecting an estimated 69 million individuals annually.[1] The true global burden remains inadequately quantified, with low- and middle-income countries (LMICs)-home to 85% of the world’s population-disproportionately affected. The Lancet Neurology Commissions on TBI (2017, 2022) emphasise that brain injury should be understood not as an isolated acute event but as a long-term, multifaceted disease process requiring sustained ‘systems of care’.[2,3]

Effective TBI management demands a comprehensive, coordinated approach spanning prevention, acute treatment, rehabilitation, and long-term follow-up. Yet significant gaps persist across centres, regions, and health systems. Care can be particularly fragmented in the post-acute phase, where resource allocation often favours acute intervention over rehabilitation and long-term follow-up.[2,4]

Digital health technologies offer promising solutions to address these gaps. The World Health Organisation (WHO) Global Strategy on Digital Health advocates leveraging digital tools within existing infrastructure to streamline care delivery, strengthen provider-patient communication, and improve data utilisation.[5]

Relevant modalities include electronic health records (EHRs), telemedicine, mobile health (mHealth), wearable technologies, and health information systems. Whilst the Lancet Commission identifies technology as crucial for transforming neurotrauma outcomes, practical guidance on effective global implementation remains limited. Existing digital health frameworks, including the Telehealth Service Implementation Model (TSIM) and WHO telemedicine framework, offer valuable perspectives but do not address the specific contextual challenges of the TBI care pathway.[6] Implementation must also account for ‘common-denominator’ technologies, such as telephone and SMS, that mitigate socioeconomic barriers while balancing data burden against clinical value. A systems thinking lens is particularly valuable in this context, recognising TBI care not as a series of discrete clinical encounters but as an interconnected system in which technological, human, organisational, and contextual factors interact across the care continuum.[7,8]

Despite growing recognition of digital health’s potential, practical tools to guide technology integration in TBI care remain underdeveloped. Clinician perspectives on contextual challenges are recognised as critical to successful implementation, yet work specific to digital technology integration across the TBI care continuum is lacking.[9] A recent scoping review of digital mental health interventions and patient-reported outcome measures in TBI populations identified a predominance of high-income country evidence and called for a systems-based approach to technology integration across the TBI care pathway-highlighting the need for the clinician-centred, global framework developed in the present study.[10,11]

This study addressed these gaps by exploring neurosurgical clinicians’ experiences with digital technologies across the TBI care pathway in diverse global health systems. The objectives were to: (1) determine current technology use and experiences across the care continuum; (2) characterise the contextual factors shaping technology integration across diverse health systems; and (3) develop a novel conceptual framework to guide digital health integration in TBI care globally.

## Materials and Methods

### Study Design

This was a cross-sectional qualitative study situated within the critical realist paradigm, which acknowledges an independently existing reality whilst recognising that understanding is mediated by context, culture, and individual experience.[12] This paradigm enabled exploration of the mechanisms underlying participant responses whilst acknowledging the interpretive nature of the research process. A systems thinking approach, guided by the Engineering Better Care (EBC) framework, informed iterative development of the semi-structured interview schedule.[13] Systems thinking focuses on understanding how components of a complex system interact and influence one another, recognising that changes in one part of a system can have cascading effects across the whole. This approach is particularly suited to TBI care, where clinical, technological, organisational, and contextual factors are deeply interconnected.[8] Published in 2017 through a partnership between the Royal Academy of Engineering, the Academy of Medical Sciences, and the Royal College of Physicians, EBC operationalises this systems approach through four central themes-people, systems, design, and risk-developed with stakeholders across clinical medicine, engineering, design, and systems science. The research question and study design were further informed by community engagement and involvement activities-including a stakeholder workshop conducted in Uganda as part of the wider NIHR ABSI research programme-which confirmed the clinical relevance of digital technology integration as a priority for investigation.

Methods are reported in accordance with the Consolidated Criteria for Reporting Qualitative Research (COREQ) guidelines; the completed checklist is provided in Supplementary Materials.[14]

### Ethics

Ethical approval was obtained from the University of Cambridge Engineering Department Research Ethics Committee (Project ID: #431). All data were stored on General Data Protection Regulation (GDPR)-compliant servers with participants assigned unique identifiers. Informed consent was obtained electronically; participants were advised that anonymised quotes would be published and that, despite anonymisation, contributions might remain identifiable to those familiar with their work or context.

### Reflexivity

The first author is a white, female medical student from the UK, whose training has predominantly engaged with positivist, quantitative methodologies. This research represented a transition into qualitative inquiry, and a background in medicine and academic interest in neurosurgery and digital health may have shaped interpretive decisions-particularly in latent coding of technology-oriented content. Conducting interviews in English, within a knowledge base largely rooted in Global North perspectives, and in a field historically dominated by such perspectives, introduced acknowledged limitations to the worldview. A reflexive journal was maintained throughout data collection and analysis, with entries documenting evolving interpretations, potential biases, and supervisory discussions. Interpretive decisions were critically reviewed with the wider research team throughout the analytical process.

### Participants and Recruitment

Inclusion and exclusion criteria for the study are outlined below in Table 1.

**Table 1:**
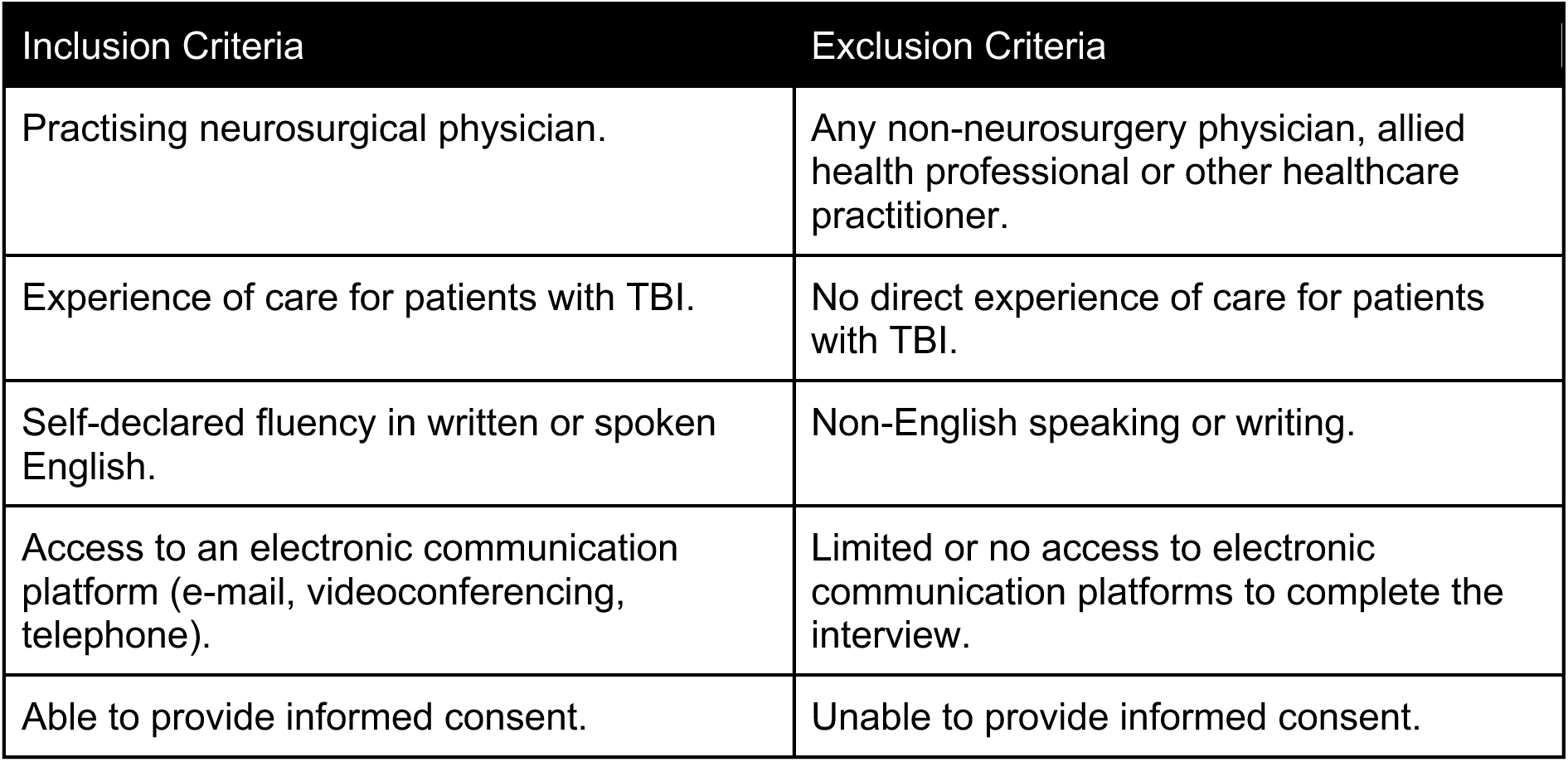
Study eligibility criteria.

| Table 1: Study eligibility criteria. |  |
| --- | --- |
| Inclusion Criteria | Exclusion Criteria |
| Practising neurosurgical physician. | Any non-neurosurgery physician, allied health professional or other healthcare practitioner. |
| Experience of care for patients with TBI. | No direct experience of care for patients with TBI. |
| Self-declared fluency in written or spoken English. | Non-English speaking or writing. |
| Access to an electronic communication platform (e-mail, videoconferencing, telephone). | Limited or no access to electronic communication platforms to complete the interview. |
| Able to provide informed consent. | Unable to provide informed consent. |

Practicing neurosurgical clinicians were recruited internationally through purposive sampling via NIHR ABSI collaborating centres, social media (Twitter, LinkedIn, WhatsApp), and the World Federation of Neurosurgical Societies (WFNS) directory newsletters. Recruitment ran throughout the data collection period (March-July 2024). All interviews were conducted by the first author and no prior relationship existed between the first author and participants before study commencement, and no non-participants were present during interviews.

Individuals expressing interest were sent a Participant Information Sheet and electronic consent form via Qualtrics™ (https://cambridge.eu.qualtrics.com). A single follow-up was sent after one week of non-response; no further contact was initiated thereafter to preserve voluntary participation. No financial or authorship incentives were offered. A formal response rate could not be calculated given the multi-channel recruitment approach.

Sample size was guided by information power principles, recognising that qualitative studies with a narrow, well-defined aim, specific participant group, and theoretically informed analysis require fewer participants to achieve sufficient information power than broader exploratory studies.[15] The aim of this study was to explore digital technology integration across the TBI care pathway from the perspective of practising neurosurgeons. The participant group was homogeneous in professional roles whilst deliberately heterogeneous in geographic and economic contexts. Data analysis occurred alongside data collection, with information power principles considered iteratively alongside coding procedures throughout recruitment. Sufficient information power was achieved after approximately 10 interviews. Additional interviews were conducted to ensure broader coverage across income settings, with broad themes identified after 12-13 interviews, and final interviews generating few new codes. Whilst information power principles do not guarantee saturation within every subgroup, the breadth of income settings spanning lower-middle, upper-middle, and high-income contexts, was considered sufficient to identify patterns of convergence and divergence across contexts relevant to the study aims.

### Data Collection

Individual semi-structured interviews were conducted in English via Microsoft Teams (March-July 2024). All interviews were video recorded and auto transcribed by Microsoft Teams; transcripts were manually corrected against audio recordings by the first author. Most interviews lasted for approximately 40 minutes, and no repeat interviews occurred. The interview schedule explored technology availability, healthcare infrastructure, clinical pathways, and contextual challenges across the TBI care continuum, with flexibility for participants to introduce relevant topics. The interview schedule was developed a priori using the four domains of the EBC framework: people, systems, design, and risk.[13] It was iteratively refined throughout data collection in response to emerging themes, participant-introduced topics, and areas of discovery relevant to the research question. The full interview guide is provided in Supplementary Materials. Demographic data, including country of residence, country of training, and practice setting, were collected verbally at the interview.

### Data Analysis

Analysis followed Wiltshire and Ronkainen’s five-step critical realist thematic analysis, conducted using NVivo™ 14.[16] The analysis was inductive, operating predominantly at an interpretive level to identify underlying mechanisms shaping participant responses rather than cataloguing surface content. Both semantic and latent coding approaches were employed, with semantic coding used for culturally sensitive concepts and latent coding applied to technology-oriented discussions where implicit meanings could be more confidently interpreted. The five steps comprised: (1) data familiarisation through iterative audio comparison and initial noting; (2) systematic coding of the full dataset generating 86 initial codes, combining semantic and latent approaches; (3) iterative theme development with supervisory input and reflexive journaling; (4) theme review and refinement-examining relationships to underlying causal mechanisms; and (5) final theme definition and naming to produce a coherent analytical narrative. Data collection and analysis proceeded in parallel. No formal member checking was undertaken; however, analytical rigour was supported by actively seeking data that challenged emerging themes. For example, returning to transcripts where participant responses did not fit developing codes, ensuring the analysis was not confirmatory in approach.

## Results

Between March and July 2024, 14 neurosurgeons from 12 countries completed semi-structured interviews (Table 2). Of 27 individuals who expressed initial interest, 14 participated; non-engagement following initial contact reflected the voluntary nature of participation and pragmatic recruitment constraints.

**Table 2:** Participant characteristics.

| World Bank Income Classification | Countries Represented (n=12) |
| --- | --- |
| Low | 0 |
| Lower-middle | 6 |
| Upper-middle | 2 |
| High | 4 |

| Country of Residence |  |  |  |
| --- | --- | --- | --- |
| Belgium | 1 | Malaysia | 1 |
| Benin | 1 | Nepal | 1 |
| Brazil | 1 | Pakistan | 1 |
| Egypt | 1 | Tanzania | 1 |
| India | 3 | United Kingdom | 1 |
| Ireland | 1 | United States of America | 1 |
| **Sex** |  |  |  |
| Male |  | 11 |  |
| Female |  | 3 |  |

Six inductive themes (Table 3) were identified from the data: 1) *Availability* 2) *Acceptability* 3) *Applicability* 4) *Capability* 5) *Feasibility* 6) *Possibility*. These have been elaborated through their related subthemes. Relevant excerpts from the interviews are integrated throughout the results section. These quotations appear in italics, followed by participant identifiers in square brackets. These quotes have been redacted as appropriate from their full verbatim transcripts (an example is evidenced in Supplementary Materials).

**Table 3:**
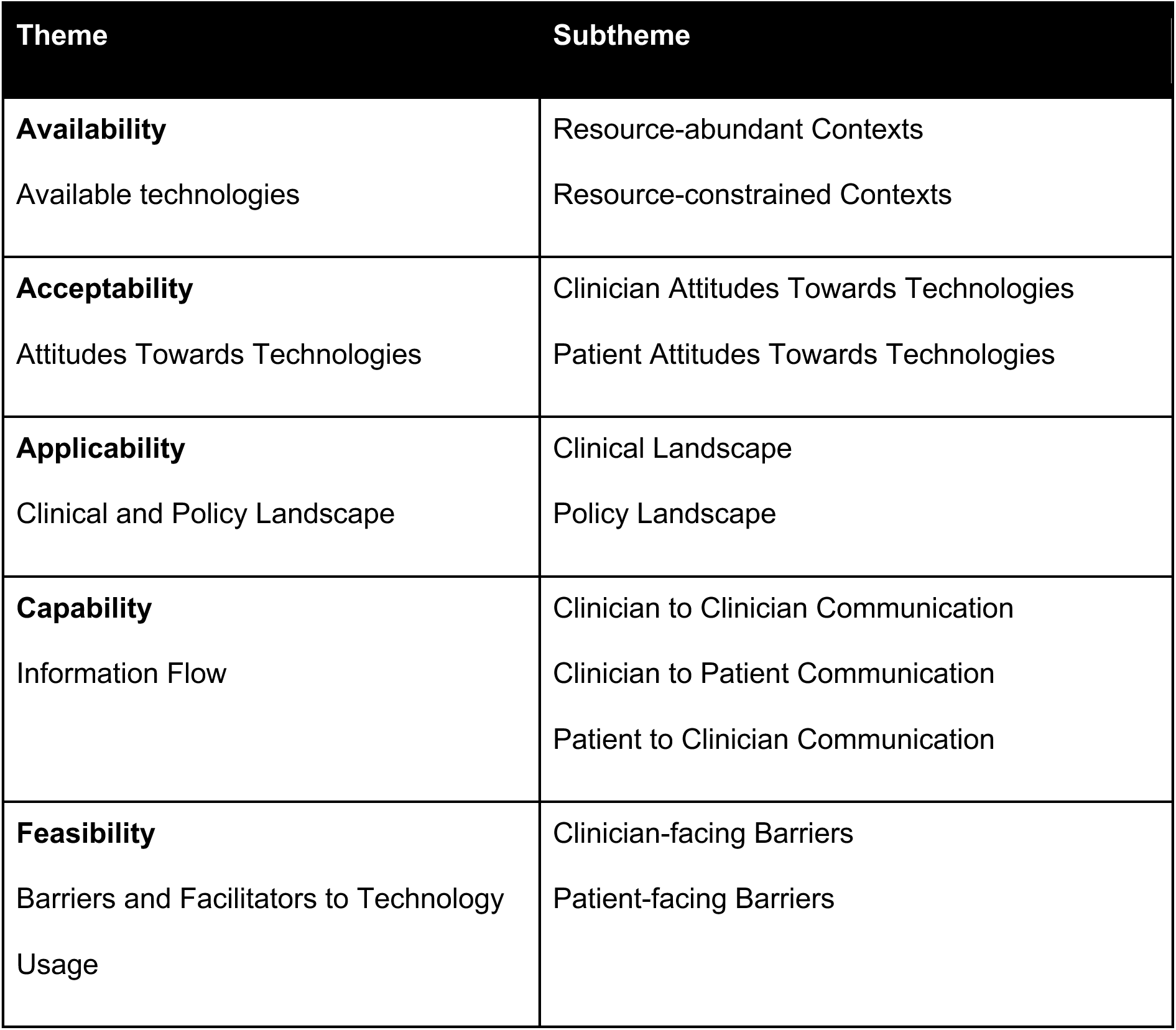
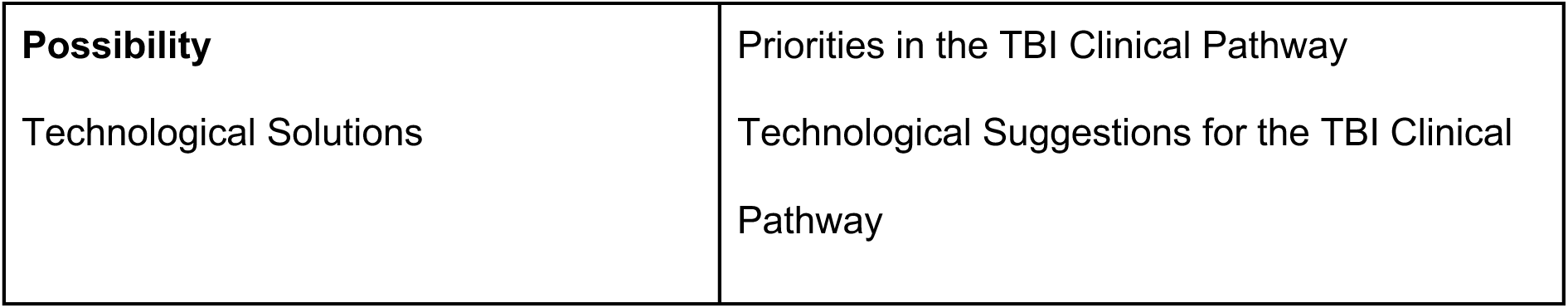
Themes and sub-themes identified from the data.

### Theme 1: Availability

Technology availability varied substantially across settings, shaped by income level, geography, and institutional infrastructure. Two distinct contexts emerged: resource-abundant and resource-constrained, though in practice many settings operated along a continuum between two ends of the spectrum.

### Resource-abundant contexts

Participants in high-income settings described access to integrated, multi-platform digital ecosystems. Electronic health record systems such as Epic-a platform integrating clinical notes, laboratory results, imaging, and prescribing into a single interface-were accessible remotely via virtual private network (VPN) and mobile applications and dedicated internal communication devices operated on closed hospital networks. Informal workarounds were common despite well-developed institutional infrastructure:

> *“We’ve had to use photographs of scans or videos of brain scans on call through WhatsApp all the time.”* [P3]

Consumer-grade technologies appeared to address specific gaps in institutional provision, particularly around the speed and accessibility of clinical information sharing in time-critical scenarios rather than representing a failure of digital infrastructure in these settings.

### Resource-constrained contexts

In lower-middle and upper-middle income settings, mobile phones-particularly smartphones-emerged as the dominant and often sole technological resource, with penetration extending into economically disadvantaged communities:

> *“Phone access is very, very omnipresent now, because the Internet is very cheap, so even the people who come below the poverty line… are able to afford cheaper smartphones.”* [P8]

Growing adoption of digital payment systems in some settings further signals expanding technological familiarity within the general population, independent of healthcare infrastructure-a potentially important facilitator for future digital health implementation.

However, a persistent urban-rural divide shaped the reliability of this access. Rural participants reported dependence on basic voice calls and short message service (SMS), with network coverage inconsistent beyond urban centres. Cost emerged as a structural barrier to digital communication more broadly, with implications for both clinician workflows and patient follow-up:

> *“Sometimes in my country, sending text messages is more expensive compared to sending WhatsApp.”* [P5]

Across both contexts, hybrid paper-digital systems were common, with clinical information flowing between printed and electronic formats depending on institutional capacity, reflecting the gap between institutional ambition and operational reality:

> *“The hospital doctor will send us the data in printed form, and we will keep that in printed form. That’s why it’s a hybrid system.”* [P11]

Grassroots intermediaries emerged in some resource-constrained settings to bridge digital literacy gaps, with community members offering paid phone and internet assistance. This enabled access to telemedical consultations where individual capability was limited [P10]. This adaptive behaviour highlights both the demand for digital health engagement and the structural inequities that necessitate such workarounds.

### Theme 2: Acceptability

Attitudes towards digital health technologies were broadly positive among clinicians, though acceptance was consistently tempered by practical and ethical concerns. Patient attitudes, reported through clinician perspectives, reflected a similar pragmatic acceptance shaped by context-specific constraints.

### Clinician attitudes

Participants across income settings valued technology primarily for its capacity to enhance communication efficiency and extend care reach, with mobile devices and messaging applications widely regarded as indispensable for urgent clinical communication:

> *“Communication is crucial, and my phone plays a key role in that. I use it to quickly reach out to team members via calls or messaging apps, especially in urgent situations.”* [P13]

However, positive attitudes were balanced by awareness of the burden technology imposes. Clinicians acknowledged the tension between technology’s long-term efficiency and the immediate demands of learning, managing, and maintaining multiple platforms:

> *“I think it’s good to have lots of communication tools but obviously sometimes it is hard to keep track.”* [P14]

A consistent view across participants was that technology should complement rather than replace clinical examination-particularly relevant in TBI care where neurological assessment relies on in-person evaluation.

### Patient attitudes

Clinicians broadly perceived patients as willing to engage with technology when they understood its clinical purpose:

> *“They know that this data is being used to help them with their clinical condition… in general the majority have no problems allowing the clinicians to use technology to improve their patient care.”* [P5]

Acceptance was nonetheless contingent on perceived safety and accessibility. Data privacy emerged as a particular concern in contexts where device-sharing is common:

> *“I always think of data security especially as some people share phones and share SIM cards too.”* [P7]

This finding underscores that patient acceptance cannot be assumed from access alone. Trust, transparency around data use, and awareness of local device-sharing practices are prerequisites for meaningful engagement. Clinicians emphasised that SMS-based communication, requiring neither smartphones nor internet connectivity, offered a pragmatic solution for diverse patient demographics including older adults and those with TBI-related functional impairment.

### Theme 3: Applicability

The applicability of digital technologies in TBI care was shaped by the clinical and policy contexts in which they must operate. These contexts varied substantially across settings, creating both opportunities and constraints for technology integration.

### Clinical landscape

Participants described a TBI care pathway characterised by fragmentation, resource strain, and significant urban-rural disparities. While urban centres reported improvements in care organisation and outcomes, rural and remote areas faced compounding challenges across the entire care continuum-from pre-hospital response through to long-term follow-up:

> *“Those patients… delays happen because they do not have any system… sometimes because they don’t know how to shift the patient, they can add more injuries to that patient.”* [P1]

Resource strain was a recurring concern, with many institutions operating well beyond capacity:

> *“The current bed strength allotted to neurosurgery in my hospital… is only 88 but at the time we have around 2 1⁄2 times the number of patients.”* [P8]

Loss to follow-up emerged as a consistent challenge across all settings, representing a critical gap in the care continuum where digital technologies could have significant impact:

> *“Quite a number of them don’t come back to hospital once they are well… they think it’s a hassle to come to hospital so quite a number of patients are lost to follow up.”* [P5]

### Policy landscape

In high-income countries, data protection legislation posed tangible barriers to informal communication. In the United States, the Health Insurance Portability and Accountability Act of 1996 (HIPAA) was cited as a particular constraint, given the breadth of what constitutes protected patient information:

> *“The biggest barrier to text messaging in our context would be like HIPAA violation… what we consider protected patient information is very, very broad.”* [P2]

Yet in practice, clinicians in multiple settings reported using consumer applications outside formal regulatory frameworks to meet patient needs. Concerns around the General Data Protection Regulation (GDPR) were similarly acknowledged:

> *“It does cross a few boundaries… WhatsApp is not encrypted… there are GDPR issues there.”* [P3]

Cultural attitudes towards healthcare providers added a further layer of complexity to the policy landscape. In some settings, high levels of deference to medical authority shaped how informed consent and patient autonomy were approached in practice [P7]. Conversely, public trust in healthcare systems was variable, with urban populations generally reporting greater confidence than rural or lower-income communities [P13]. Trust in healthcare systems varied considerably across settings, and this variation has practical consequences for how digital health technologies are designed and implemented.

### Theme 4: Capability

Capability describes the information flow across the TBI care ecosystem, examining how effectively communication occurs between clinicians, and between clinicians and patients. Across all settings, a tension emerged between the communication tools available and those actually used in practice.

### Clinician-to-clinician communication

Institutional communication infrastructure was frequently inadequate, leading clinicians to rely on personal devices and consumer applications for work-related communication:

> *“I personally use my personal phone to call my colleague, to call the patient… because the hospital did not give us the phone.”* [P10]

Messaging applications such as WhatsApp were widely used for rapid sharing of clinical information and imaging, particularly in settings without comprehensive electronic records. Where secure integrated platforms existed, adoption was often limited by access barriers:

> *“We also have a secure platform that’s integrated into our electronic medical record but I would say this is not as common as you can’t access from home unless you have gone through the long sign up.”* [P14]

This pattern-formal systems underutilised in favour of informal workarounds-was consistent across income settings, reflecting a gap between institutional provision and clinical workflow needs. Hybrid paper-digital systems remained common, particularly in lower-middle-income settings [P11], reinforcing findings from Theme 1.

### Clinician-to-patient communication

Communication with patients was characterised by adaptability, with clinicians employing a range of methods according to patient capability and access. Where advanced technologies were unavailable or impractical, clinicians defaulted to telephone calls and illustrated written instructions for family caregivers [P12, P13]. Video conferencing, despite its potential, was underutilised:

> *“We don’t tend to use video conferencing with patients… our computers in the hospital are so rubbish that it’s just not worth the hassle.”* [P3]

This assessment highlights how infrastructure quality, not just availability, determines whether technologies are adopted in practice.

### Patient-to-clinician communication

Where patient portal systems existed, they fundamentally altered communication dynamics but introduced new burdens for clinicians:

> *“Now suddenly we’ve added this expectation that you’re going to be answering essentially constant messages… they don’t go away during vacation or weekends.”* [P2]

This ‘always on’ expectation raises important considerations for technology design-particularly the need to build in triage and boundary mechanisms to prevent clinician burden from undermining sustained adoption. Professional boundary management emerged as a related challenge, with clinicians receiving unsolicited patient contact via personal numbers [P1], further blurring the distinction between institutional and informal communication channels.

### Theme 5: Feasibility

Feasibility examines the practical barriers encountered by both clinicians and patients in adopting and sustaining digital health technologies across the TBI care pathway. Barriers were interconnected and often mutually reinforcing, reflecting the systemic nature of implementation challenges across global contexts.

### Clinician-facing barriers

Resource scarcity emerged as a foundational barrier, particularly in lower-middle-income settings, constraining not only access to technologies but the trained personnel required to operate and maintain them:

> *“We are working in a very, very difficult setting because we lack a lot of things.”* [P10]

The inherent limitations of remote assessment in TBI care presented a further challenge. Participants acknowledged telemedicine’s potential whilst recognising its inability to replicate the neurological examination:

> *“Obviously I believe telemedicine is not the right answer, but sometimes you are not able to have that much of a good evaluation virtually.”* [P11]

Infrastructural instability, particularly unreliable internet connectivity, undermined confidence in technology-dependent care pathways:

> *“I think that we should also mention something about the internet interruption that really happens if we are going to be making everything virtual.”* [P11]

Increased clinician workload and the always-on expectations introduced by digital communication platforms, raised in Theme 4, were identified as sustainability concerns, with potential for burnout and long-term adoption. Nonetheless, facilitators to implementation were also evident: clinician adaptability, widespread adoption of consumer-grade communication tools, and grassroots community mechanisms extending digital access beyond formal infrastructure-findings elaborated across Themes 1, 2, and 4-demonstrated that implementation capacity exists even where institutional provision falls short.

### Patient-facing barriers

Financial constraints posed a fundamental barrier to sustained digital engagement, particularly in prepaid mobile markets:

> *“Most of our SIM cards are prepaid… sometimes they can’t afford to pay, and then they go incommunicado. And if they don’t pay for three months, that number goes to somebody else.”* [P7]

This finding has direct implications for both continuity of care and patient confidentiality. A disconnected number does not simply represent lost contact, but a patient potentially lost to follow-up entirely, whilst number reassignment introduces a risk of sensitive clinical information reaching an unintended recipient. The digital divide manifested across multiple dimensions: device capability, network reliability, and digital literacy, with urban-rural disparities persisting across income settings [P13]. Geographic barriers compounded these challenges in rural areas, where terrain and distance affected both physical access to care and telecommunications infrastructure [P13].

Language and cultural complexity introduced additional barriers to text-based communication in multilingual settings:

> *“It makes counselling sometimes interesting because you swap from language to language… so texting would be very hard, I mean I might not know three of the four languages I need.”* [P7]

Age-related barriers were significant in high-income settings, where older patients struggled to engage with sophisticated patient portal systems despite willingness to use simpler technologies such as telephone and SMS [P2]. This finding reinforces the case for common-denominator technologies as a pragmatic baseline for inclusive digital health design.

### Theme 6: Possibility

Possibility synthesises clinician visions for future digital health integration across the TBI care pathway, identifying both priority areas for intervention and specific technological solutions. Collectively, these perspectives informed the development of the conceptual framework presented from this study.

### Priorities in the TBI clinical pathway

Participants consistently identified the post-discharge period as the most critical and underserved point in the care continuum from a technological standpoint, recognising that whilst pre-hospital and acute vulnerabilities are substantial, the interval between discharge and follow-up is where digital solutions are most lacking, and patients have least contact with their care team:

> *“I think that the technology will be better between the discharge and the follow up… if we could have a technology where we get to follow up and know our morbidity and mortality in the long-term outcome of our patients.”* [P9]

Remote monitoring and virtual follow-up were identified as priority solutions for this gap, with direct patient-to-team communication channels seen as essential:

> *“I think one of the most important areas is around remote monitoring and remote care. Video consultations for follow-up visits and creating more opportunities for patients to directly communicate with the care team. Because there is this big gap.”* [P14]

Improving intra- and inter-hospital communication emerged as an acute care priority, with participants expressing frustration at existing switchboard-dependent systems. Enhanced referral pathways and real-time pre-hospital data sharing were highlighted as opportunities to improve acute outcomes, particularly for rural patients:

> *“If there were more portable ways to provide care, that would be good, and also ways to track ambulances and give real-time data to the hospital about who’s coming so they can better prepare.”* [P13]

Standardised data collection and reporting across the care continuum was identified as a systemic priority, underpinning all other improvements:

> *“Reporting is problematic. There’s no homogenised reporting.”* [P4]

This absence of standardisation limits outcome tracking, research capacity, and cross-institutional learning across all income settings. Without consistent reporting measures, data from a neurosurgical unit in a lower-middle-income setting cannot be meaningfully compared with or contribute to international registries, limiting the global evidence base for TBI care improvement.

### Technological suggestions

Clinicians proposed solutions spanning the full care continuum. Telemedicine and video conferencing infrastructure were consistently highlighted for connecting peripheral centres with specialist facilities [P11]. Integrated trauma registries capable of federating data across institutions and settings were identified as foundational requirements:

> *“We’re obviously looking at trauma registries collecting all that data and federating all the data, linking it all up in one place and trying to work out a way that’s secure.”* [P4]

Real-time ambulance tracking and portable diagnostic technologies were proposed for the acute phase [P13], whilst patient-facing applications supporting self-monitoring and symptom reporting were envisioned for the post-discharge period. Across all suggestions, participants acknowledged the tension between technological ambition and equitable access:

> *“We need to find the balance to make sure we don’t leave the people in the rural country behind.”* [P14]

Ensuring that digital health innovation does not deepen existing disparities runs as a unifying thread across all six themes and sits at the heart of the framework developed from this study.

## Discussion

To our knowledge, this is the first qualitative study to describe neurosurgical clinicians’ perspectives on digital technology integration across the TBI care pathway in diverse global settings.[17] Six inductive themes - Availability, Acceptability, Applicability, Capability, Feasibility, and Possibility - revealed an interplay between technological potential and the multifaceted challenges of healthcare delivery. Together these formed the basis of a novel conceptual framework to guide digital health technology design and implementation in TBI care globally.[4]

The disparities in technology availability and utilisation identified across income settings and urban-rural divides reflect broader patterns documented in the digital health literature.[18] Whilst smartphone penetration has increased substantially across LMICs, connectivity reliability, digital literacy, and affordability remain persistent barriers to consistent engagement with digital health tools.[19] Our findings extend this literature by demonstrating that these barriers operate simultaneously and interact with one another. Financial constraints limiting prepaid mobile access, for instance, directly undermine continuity of follow-up care in ways that infrastructure investment alone cannot resolve.

The widespread adoption of consumer-grade technologies, particularly WhatsApp and SMS as clinical workarounds, reflects adaptation documented across multiple health contexts, [18] yet raises unresolved tensions around data governance and regulatory compliance.[20,21] Clinicians reported knowingly operating outside GDPR and HIPAA frameworks to meet patient needs. This regulatory versus reality gap represents a critical design challenge: technologies developed for high-income regulatory environments are poorly suited to the contexts in which they are most needed.

The emergence of grassroots intermediaries, such as community members offering paid assistance with phone access and internet connectivity to enable telemedical consultations for those with limited capability, is a finding with implications beyond TBI care. These informal infrastructural elements-including shared device access and community-based digital literacy support-operate entirely outside formal health systems yet meaningfully extend their reach. These findings suggest that community-level facilitators can be as important as institutional provision in determining whether digital health technologies reach those who need them most, and that implementation strategies must account for this informal layer of infrastructure.

### The novel conceptual framework

The six themes identified in this study-Availability, Acceptability, Applicability, Capability, Feasibility, and Possibility-collectively constitute a conceptual framework for understanding digital health integration in TBI care globally. Rather than emerging from a single pre-existing model or a Best-Fit Framework Synthesis, this framework was derived from the iterative synthesis of primary qualitative data, reflexive practice, and community engagement and involvement activities conducted as part of the wider NIHR ABSI research programme. The themes are arranged across two overarching constructs: Intervention and Adoption, encompassing Availability, Capability, and Possibility; and Context and Infrastructure, encompassing Applicability, Acceptability, and Feasibility. These emerged from the analytical process of creating themes that describe the characteristics and uptake of technologies from those describing the contextual and infrastructural conditions in which they must operate. As a conceptual tool, the framework gives researchers, clinicians, and policymakers a consistent set of considerations to think through when planning digital health implementation in any given context, without prescribing what the answer should be.

The hexagonal form was selected as a communication device (Fig 1). and was chosen for its capacity to make the full range of contextual considerations visible simultaneously at a glance, rather than to imply scalar measurement. The chart has no numerical axes and the shaded areas in the example scenarios are illustrative of relative emphasis rather than precise scores. It does not function as a quantitative scoring instrument or an implementation science framework prescribing how technologies should be deployed. Rather, it operates as a structured guide for contextual thinking and prompts consideration of the six dimensions that shape whether a digital health technology is likely to work in each setting. The themes were not derived with the assumption that any one carries universal priority over another. Instead, the framework is designed to surface whichever dimensions are most constraining or most enabling in a specific context. Where infrastructure is severely limited, greater emphasis may be on Availability and Feasibility. Conversely, where infrastructure is adequate, Acceptability and Capability may become more critical considerations.

**Figure 1:**
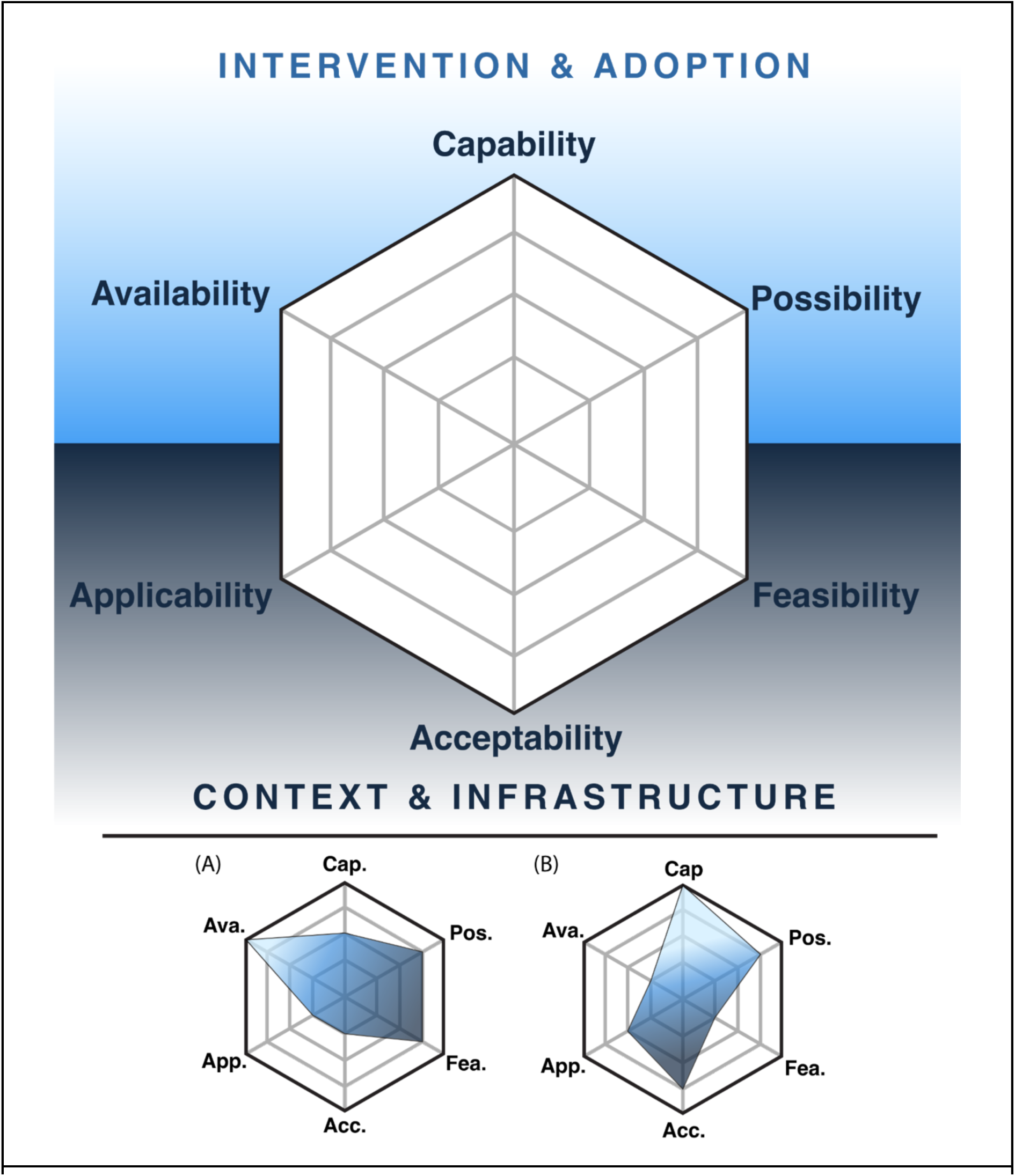
A novel conceptual framework for digital health in the TBI care pathway The framework represents six inductive themes arranged across two conceptual domains: Intervention and Adoption (Availability, Capability, Possibility) and Context and Infrastructure (Applicability, Acceptability, Feasibility). Example scenarios illustrate how different technologies might be considered using the framework across the six dimensions: (A) SMS follow-up. This is accessible across basic phones without internet connectivity and low-cost to implement but limited in the richness of clinical information they can transmit or collect. (B) Video telemedicine platform with integrated patient records-clinically powerful and innovative, but requiring reliable internet, compatible devices, and institutional infrastructure that limits its reach in resource-constrained settings. Ava. = Availability; App. = Applicability; Cap. = Capability; Acc. = Acceptability; Pos. = Possibility; Fea. = Feasibility.

Existing implementation frameworks, including TSIM,[6] the WHO telemedicine framework,[21] and the IDEAL Collaborative,[22] provide process-oriented and organisational perspectives on how digital health technologies and surgical innovations can be systematically developed and deployed. Similarly, the DH-EquIR model [23] offers a structured approach to equity-focused digital health implementation across five phases: population health, planning, designing, implementing, and measuring equitable outcomes. These frameworks are predominantly process focused. They describe how to move through stages of development and assessment, but do not provide the contextual specificity required to understand whether a given technology is appropriate for a chosen setting before or during that process.

Rather than describing implementation stages, this framework provides specific contextual criteria that designers, clinicians, and policymakers can use to map the implementation landscape. To illustrate: a team developing a telemedicine follow-up platform for TBI patients in a particular context might use IDEAL to structure their staged development process and DH-EquIR to ensure equity considerations are embedded at each phase. The present framework could be applied alongside these to understand whether the digital infrastructure exists to support the platform (Availability), whether clinicians and patients in that context are likely to adopt it (Acceptability), and whether the regulatory environment permits its use (Applicability). These contextual criteria directly inform the design decisions made at each stage of both models. Used together, the frameworks provide both a process roadmap and a contextual map.

### Strengths and Limitations

This study’s international scope, spanning twelve countries across three World Bank income classifications, provides cross-contextual insights rarely achieved in qualitative neurotrauma research.[24] The use of critical realist thematic analysis, with reflexive journaling and supervisory review throughout, enhances the credibility and transparency of the analytical process. The resulting framework has strong internal integrity-each theme is directly traceable to participant data and quotes, and the consistent naming logic of the six domains supports both coherence and practical usability.

Limitations include the constraints of single-researcher data collection and analysis, which will inevitably have shaped interpretive decisions despite reflexive mitigation. Interviews conducted exclusively in English may have excluded perspectives from non-English-speaking neurosurgeons, potentially biasing the sample towards English-fluent practitioners and limiting representation from some LMIC contexts. Limited email response rates within a one-year timeframe may have further introduced selection bias towards those with greater engagement with international neurosurgical networks. Notably, no participants from low-income countries were recruited, limiting the framework’s direct applicability to the most resource-constrained settings and representing a priority for future work. The sample of fourteen participants, whilst appropriate for the aims of qualitative inquiry and consistent with information power principles, does not capture the full heterogeneity of global neurotrauma practice. Member checking was not undertaken; however, analytical rigour was supported by actively examining data that did not fit emerging themes, ensuring the analysis was not confirmatory in approach.[14]

Consistent with qualitative research conventions, findings are not intended to be statistically generalisable but offer theoretical transferability. The framework and themes provide conceptual tools that readers can assess for relevance to their own clinical and implementation contexts, recognising that the conditions shaping applicability will vary across settings.

### Future directions

Future work should prioritise expansion of the participant sample to include a broader range of neurosurgical clinicians, allied health professionals, policymakers, and critically, patients and caregivers. Incorporating non-English-speaking contexts and a wider range of low-income country settings would address the most significant sampling limitation identified. Second coding by an independent researcher would further strengthen analytical rigour. Ultimately, prospective validation of the framework across specific digital health interventions in TBI care will be essential to establishing its utility as a practical implementation tool.

Additional work may involve assigning numerical values across the six dimensions-such as through trade-off analysis or, for example, using smartphone penetration rates as a proxy for Availability. This was outside the scope of the present qualitative study, and any such translation would require careful validation to ensure it does not imply precision the framework is not designed to convey.

### Conclusions

This study presents the first qualitative framework developed from international neurosurgical clinician perspectives to guide digital health technology integration across the TBI care pathway. The six-theme framework offers a practical, context-sensitive tool for researchers, clinicians, and policymakers navigating the complex realities of digital health implementation in diverse global settings. As investment in digital health accelerates globally, frameworks grounded in clinical experience and contextual evidence will be essential to ensuring that technological innovation translates into equitable improvements in TBI care.[25]

## Data Availability

The data underlying this study consist of qualitative interview transcripts. Due to the small sample size (14 participants across 12 countries), full transcripts cannot be shared publicly as participants may be identifiable despite anonymisation, and participants were advised of this limitation at consent. An example of a redacted, anonymised transcript is provided in the Supplementary Materials. Researchers seeking further information may contact the corresponding author.

## Supporting Information

**Supplementary Information** contains the following: S1) Semi-structured interview guide, overlaid with the four principles of the Engineering Better Care framework; S2) Wiltshire and Ronkainen five-step critical realist thematic analysis approach; S3) Redacted verbatim transcript example; S4) COREQ (Consolidated Criteria for Reporting Qualitative Research) checklist.

## Acknowledgments

PJH is supported by the NIHR (Senior Investigator Award, NIHR Global Health Research Group on Acquired Brain and Spine Injury, NIHR Health Tech Research Centre for Brain Injury, Cambridge Biomedical Research Centre) and the MRC (TBI-Reporter Grant).

## Notes

### Author Declarations

Ethical approval was granted by the University of Cambridge Engineering Department Research Ethics Committee, Project ID #431.

